# Trans-ancestry epigenome-wide association meta-analysis of DNA methylation with lifetime cannabis use

**DOI:** 10.1101/2022.12.29.22284040

**Authors:** Fang Fang, Bryan Quach, Kaitlyn G. Lawrence, Jenny van Dongen, Jesse A. Marks, Sara Lundgren, Mingkuan Lin, Veronika V. Odintsova, Ricardo Costeira, Zongli Xu, Linran Zhou, Meisha Mandal, Yujing Xia, Jacqueline M. Vink, Laura J Bierut, Miina Ollikainen, Jack A. Taylor, Jordana T. Bell, Jaakko Kaprio, Dorret I. Boomsma, Ke Xu, Dale P. Sandler, Dana B. Hancock, Eric O. Johnson

**Affiliations:** GenOmics, Bioinformatics, and Translational Research Center, RTI International, Research Triangle Park, NC, USA; Epidemiology Branch, National Institute of Environmental Health Sciences, National Institutes of Health, Research Triangle Park, NC, USA; Department of Biological Psychology, Amsterdam Public Health Research Institute, Vrije Universiteit Amsterdam, Amsterdam, The Netherlands; Institute for Molecular Medicine Finland FIMM, University of Helsinki, Helsinki, Finland; Department of Psychiatry, Yale School of Medicine, West Haven, CT, USA; Department of Twin Research & Genetic Epidemiology, King’s College London, London, UK; Behavioural Science Institute, Radboud University, Nijmegen, The Netherlands; Department of Psychiatry, Washington University in Saint Louis School of Medicine, St. Louis, Missouri, USA; VA Connecticut Healthcare System, West Haven, CT, USA; Fellow Program, RTI International, Research Triangle Park, NC, USA; Department of Psychiatry, University of Groningen, University Medical Center Groningen, Groningen, The Netherlands

## Abstract

Cannabis is widely used worldwide, yet its links to health outcomes are not fully understood. DNA methylation can serve as a mediator to link environmental exposures to health outcomes. We conducted an epigenome-wide association study (EWAS) of peripheral blood-based DNA methylation and lifetime cannabis use (ever vs. never) in a meta-analysis including 9,436 participants (7,795 European and 1,641 African ancestry) from seven cohorts. Accounting for effects of cigarette smoking, our trans-ancestry EWAS meta-analysis revealed four CpG sites significantly associated with lifetime cannabis use at a false discovery rate of 0.05 (*p* < 5.85 × 10^−7^): cg22572071 near gene *ADGRF1*, cg15280358 in *ADAM12*, cg00813162 in *ACTN1*, and cg01101459 near *LINC01132*. Additionally, our EWAS analysis in participants who never smoked cigarettes identified another epigenome-wide significant CpG site, cg14237301 annotated to *APOBR*. We used a leave-one-out approach to evaluate methylation scores constructed as a weighted sum of the significant CpGs. The best model can explain 3.79% of the variance in lifetime cannabis use. These findings unravel the DNA methylation changes associated with lifetime cannabis use that are independent of cigarette smoking and may serve as a starting point for further research on the mechanisms through which cannabis exposure impacts health outcomes.

## Introduction

Cannabis use is highly prevalent around the world.^1^ In the United States, the legal use of cannabis has expanded across the states over time.^2^ Despite the potential therapeutic benefits of medical use,^3,4^ the widespread recreational use of cannabis has raised concerns because of its reported associations with many adverse health outcomes, including mental health (anxiety, depression, psychosis, schizophrenia, and mania),^5-8^ cognitive deficits,^9-11^ and addiction.^12^ It is a pressing public health issue to better understand the full spectrum of the benefits and adverse consequences associated with cannabis use.

DNA methylation (DNAm), which involves the addition of a methyl group to the C5 position of cytosine in the context of CpG dinucleotides, has been extensively studied in relation to gene expression and can be influenced by the genome, the environment, and stochastic processes.^13-15^ The DNAm changes induced by environmental exposure are sometimes persistent and long-lasting, while others are transient and reversible. For example, cigarette smoking has been shown to induce DNAm changes at CpGs throughout the genome. Some of these DNAm changes may revert after smoking cessation, while other DNAm changes may persist for years after cessation.^16^

In recent years, research toward understanding the effect of cannabis use on DNAm has grown.^17^ Previous candidate gene studies identified DNAm changes of *CB1 receptor*^18^ and *DAT1*^19^ in cannabis-dependent patients, *COMT* in adolescents defined as high-frequent cannabis users (> four times in the past 4 weeks),^1^ and *DRD2* and *NCAM2* in moderate to heavy cannabis users (> 10 days in the last 30 days).^20^ The first epigenome-wide association study (EWAS) of cannabis use, which compared DNAm between 12 cannabis users and 12 non-users in human sperm, found at least 10% DNAm differences at 3,979 CpG sites.^21^ Our group performed the first blood-based EWAS of lifetime cannabis use (ever vs. never) in 2,583 women and found significant DNAm changes at cg15973234 (*CEMIP*).^22^ Cannabis use– associated DNAm changes in blood have also been reported in heavy cannabis users (N = 96)^23^ and adolescents (N = 525).^24^ Taken together, these studies provide evidence that cannabis use impacts the epigenome, but knowledge of specific DNAm changes remain limited.

In this study, we conducted the largest trans-ancestry EWAS meta-analysis for lifetime cannabis use (ever vs. never) in 9,436 participants from seven cohorts. The initial model, which adjusted for sex, age at blood collection, blood cell proportions, and technical covariates, yielded 608 significant (False Discovery Rate (FDR) < 0.05) CpGs, among which 82% overlapped with prior EWAS findings for cigarette smoking. We explored this finding further by first adjusting the analyses for cigarette smoking status and next conducted the EWAS in participants who never smoked cigarettes. These two analyses identified a total of five cigarette smoking–independent CpGs significantly associated with lifetime cannabis use. We evaluated these findings by constructing a methylation score, summarizing regional DNAm changes using differential methylation region (DMR) analysis, and integrating the DNAm findings with gene expression and genetic data.

## Results

### Sample characteristics

The demographic characteristics of 9,436 study participants are summarized in Table 1. The sample consisted of 57% females, and the mean age at DNAm sampling ranged from 17.1 years in the ALSPAC cohort to 58.8 years in the TwinsUK cohort. A total of 44% of the participants reported having used cannabis at some point in their lives. DNAm sites were assessed using either the Illumina HumanMethylation 450K Bead-Chip (76%) or the Illumina HumanMethylation EPIC (850K) Bead-Chip (24%). The sample included individuals of both European ancestry (EA, 83%) and African ancestry (AA, 17%).

**Table 1.** Characteristics of the cohort participants.

| **Study Cohort** | **Total N** | **Countries** | **Ancestry** | **Female %** | **Age at blood draw\*** | **DNAm array** | **Cannabis Ever users** |
| --- | --- | --- | --- | --- | --- | --- | --- |
| Sister Study | 2,583 | US | 100% EA<sup>§</sup> | 100% | 57.0±8.8 | 450K | 49% |
| GuLF | 1,195 | US | 56% EA<br>44% AA | 0% | 44.3±11.8 | EPIC | 56% |
| NTR | 2,142 | Netherland | 100% EUR | 70% | 37.4±13.2 | 450K | 26% |
| VACS | 1,116 | US | 100% AA | 0% | 48.9±7.9 | 450K,EPIC | 79% |
| FinnTwin | 1,368 | Finland | 100% EUR | 55% | 23.5±1.9 | 450K,EPIC | 27% |
| ALSPAC | 922 | UK | 100% EUR | 52% | 17.1±1.0 | 450K | 40% |
| TwinsUK | 110 | UK | 100% EUR | 100% | 58.8±9.0 | EPIC | 27% |
| *Mean and standard deviation presented; |  |  |  |  |  |  |  |
| §Abbreviations: EA: European American; EUR: European; AA: African American |  |  |  |  |  |  |  |

### EWAS meta-analysis results for lifetime cannabis use

We conducted EWAS meta-analysis on peripheral blood DNAm data from 9,436 participants for lifetime cannabis use. In each cohort, we tested the association between lifetime cannabis use and DNAm at each CpG site using either a linear model (for unrelated participants) or a generalized estimation equation (GEE) model (for related participants) with the family ID as the clustering variable. To investigate the confounding effect of cigarette smoking, we compared two models. Model 1 included sex (in cohorts with more than one sex), age at blood collection, measured or estimated white blood cell proportions, and technical covariates. In addition to these covariates, Model 2 included cigarette smoking status defined as current, former, or never. The EWAS meta-analysis with Model 1 identified 608 CpGs significantly associated with lifetime cannabis use at an FDR threshold of 5%. Of these, 500 CpGs had been previously reported as being significantly associated with cigarette smoking^25,26^ (Supplementary Fig. 1). After adjusting for cigarette smoking, Model 2 EWAS meta-analysis identified four CpGs (Fig. 1; Table 2) significantly (FDR < 0.05) associated with lifetime cannabis use. None of these four cannabis use–associated CpGs had been reported as being significant in previous EWAS for cigarette smoking after accounting for multiple testing (*p* > 0.05/4).^26^ The quantile-quantile (QQ) plots from both models suggested minimal inflation (*λ* = 1.1). Although many CpGs did not reach epigenome-wide significance with Model 2, their effect sizes showed consistent directions of associations as in Model 1 (Supplementary Fig. 2), and their effect sizes were highly correlated (*r* = 0.85). The four cigarette smoking-independent CpGs identified with Model 2—cg01101459, cg22572071, cg15280538, and cg00813162—were annotated to *LINC01132, ADGRF1, ADAM12*, and *ACTN1*, respectively, as the closest genes (Supplementary Fig. 3). The full list of top CpGs (*p* < 0.001) from the Model 2 EWAS meta-analysis are summarized in Supplementary Table 1.

**Table 2.**
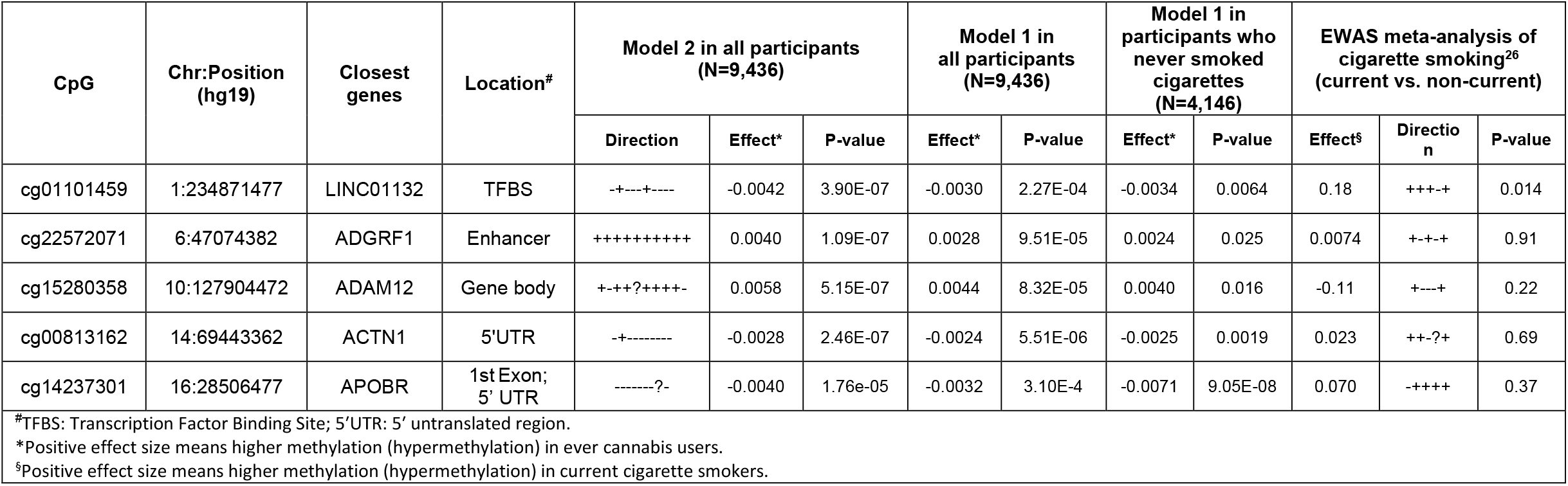
Epigenome-wide significant (FDR<0.05) CpGs identified in EWAS meta-analyses for lifetime cannabis use from Model 2, adjusted for cigarette smoking.

**Figure 1.**
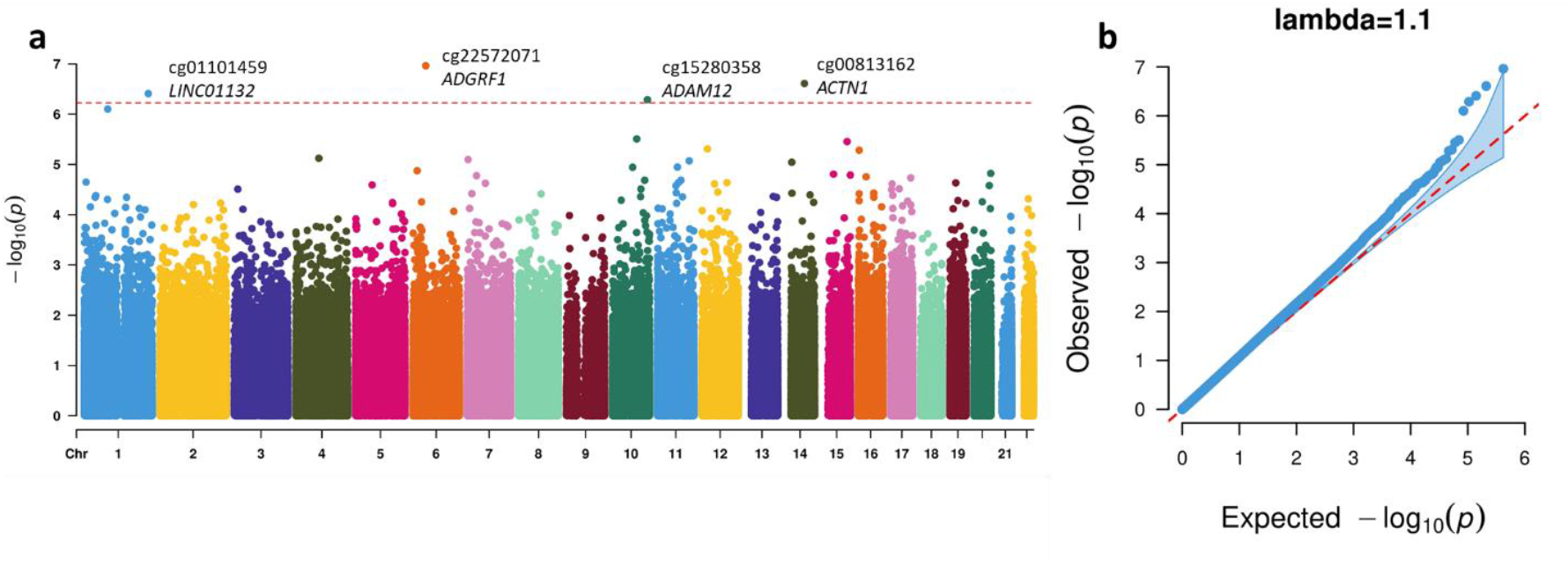
Manhattan (a) and QQ plots (b) from the EWAS meta-analysis for lifetime cannabis use with Model 2 adjusted for cigarette smoking. The dotted red line in (a) indicates the epigenome-wide significance cutoff at FDR<0.05 (*P* < 5.96 × 10^−7^).

Considering potential differences by ancestry, we performed EWAS meta-analyses stratified into EA (N = 7,795) and AA (N = 1,641) groups. The ancestry-specific results for the top CpGs reported from the Model 2 EWAS meta-analysis are shown in Supplementary Table 1. The effect sizes for the top CpGs were highly correlated between the EWAS results in the EA and AA groups (*r* = 0.77, Supplementary Fig. 4).

### EWAS meta-analysis results for lifetime cannabis use in participants who never smoked cigarettes

To further explore the confounding effect of cigarette smoking on DNAm, we conducted an EWAS meta-analysis on the subset of participants who reported never having smoked cigarettes (N = 4,146). The EWAS meta-analysis, using Model 1, identified one CpG significantly associated with lifetime cannabis use at FDR < 0.05 in this subset of participants (Fig. 2; Table 2). This CpG site, cg14237301, is annotated to the gene *APOBR*, which has been reported to be significantly associated with lifetime cannabis use in a genome-wide association study (GWAS)^27^ (Fig. 3). The full list of top CpGs (*p* < 0.001) from the EWAS meta-analysis in participants who never smoked cigarettes is summarized in Supplementary Table 2. Their effect sizes were highly correlated with the EWAS meta-analysis results in all participants under either model (*r* > 0.7, Supplementary Fig. 2).

**Figure 2.**
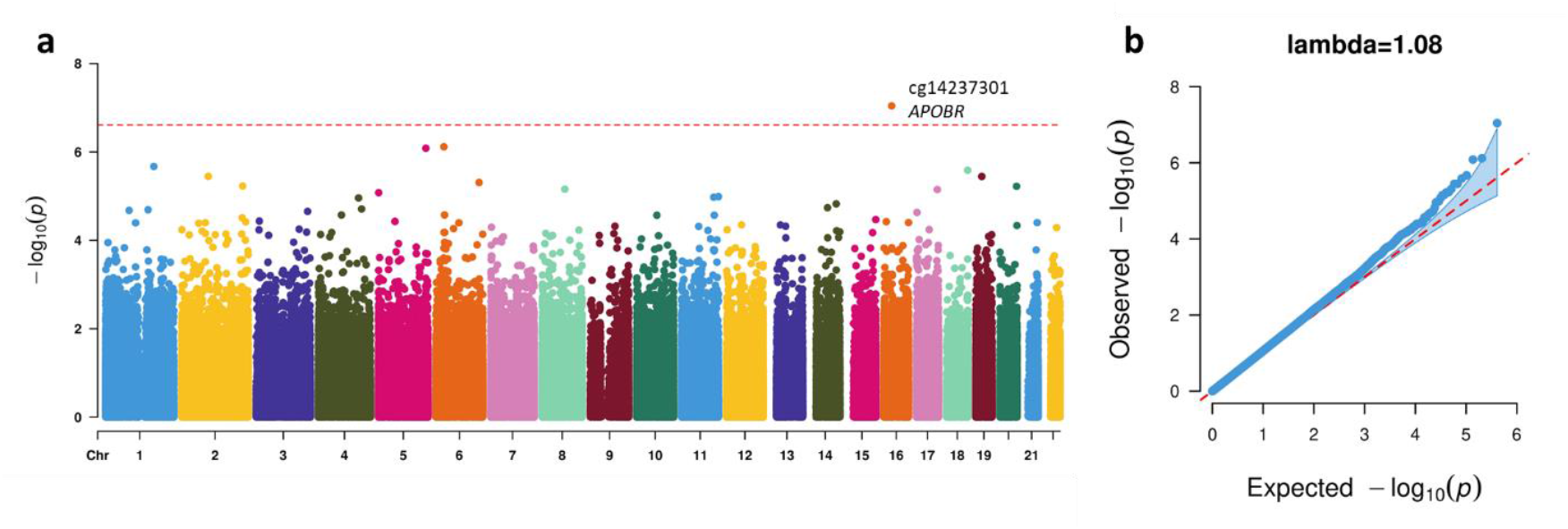
Manhattan (a) and QQ plots (b) from the EWAS meta-analysis for lifetime cannabis use in participants who never smoked cigarettes. The dotted red line in (a) indicates the epigenome-wide significance cutoff at FDR<0.05 (*P* < 2.45 × 10^−7^).

**Figure 3.**
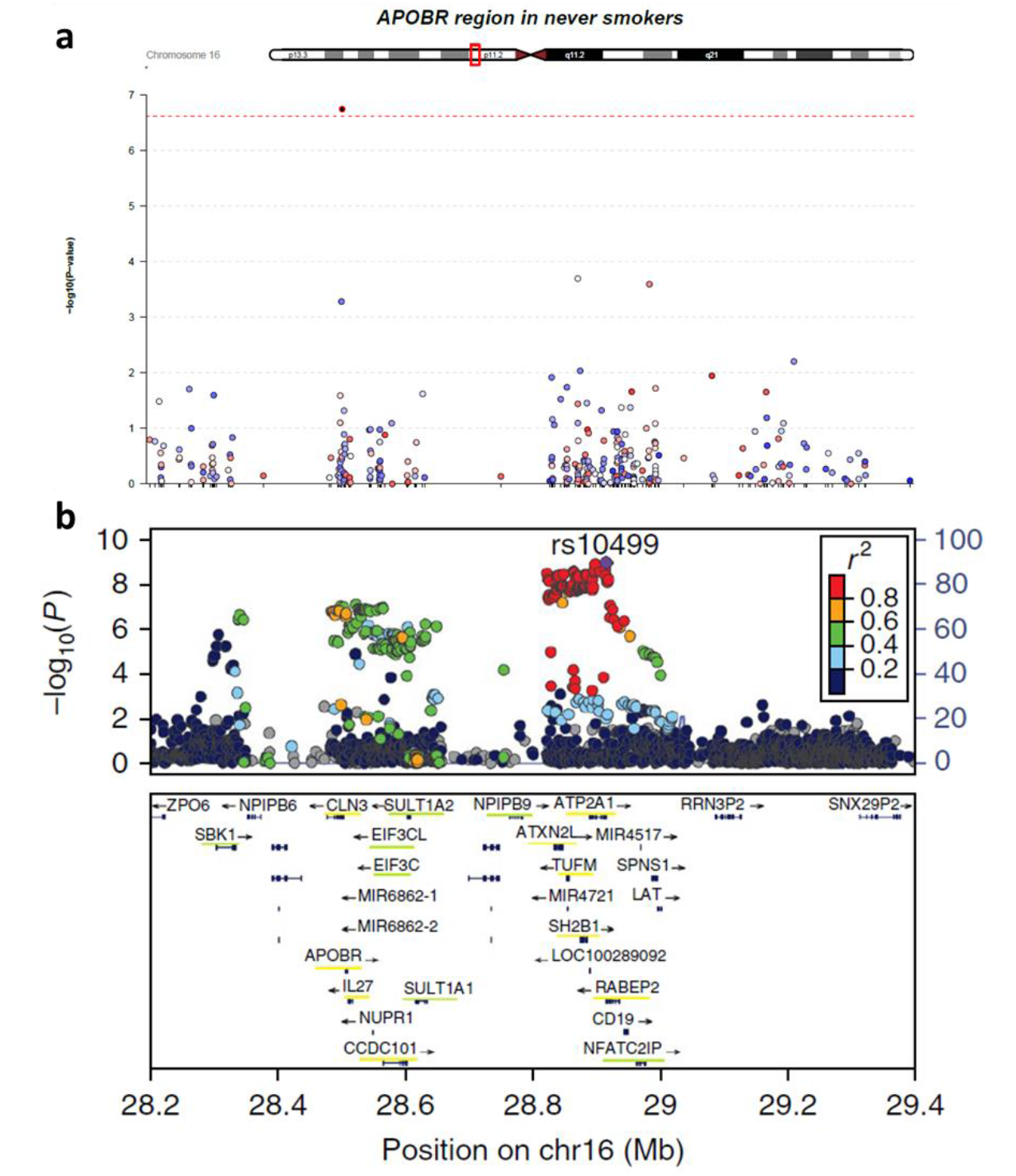
Regional plot for EWAS results in participants who never smoked cigarettes (a) and GWAS results (b) for lifetime cannabis use around the genes *APOBR/CLN3*. The x-axis shows the genomic position in base pair (bp) in hg19, while the y-axis shows the significance of associations (-log_10_ p-values). (a) Each dot is a CpG probe, and the red dotted line indicates the epigenome-wide significance at FDR<0.05. (b) Each dot is a SNP site, and the colors show different levels of LD in *r*^2^. The box in the bottom includes the genes within the genomic region, and genes underlined in yellow were significant in the gene-based test while those underlined in green were identified in the S-PrediXcan analysis^27^.

### Methylation scores

To assess the ability of DNAm levels to predict lifetime cannabis use, we calculated a methylation score based on summary statistics from the EWAS meta-analyses without the Sister Study, the largest cohort in this project. We then compared the performance of multiple models with different p-value cutoffs by the variance in lifetime cannabis use explained by the methylation scores (Supplementary Table 3). The best-performing methylation score, based on 50 CpGs with *p* < 10^−9^ in the EWAS meta-analysis using Model 1, explained 3.79% of the variance of lifetime cannabis use in the Sister Study (*p* = 1.71 × 10^−17^). To further examine the confounding effect of cigarette smoking, we also evaluated the variance explained by the methylation score in participants who never smoked cigarettes. In this subset of participants, the methylation score based on the same 50 CpGs explained 0.91% of the variance of lifetime cannabis use (*p* = 1.19 × 10^−3^). In contrast, the methylation score constructed based on EWAS meta-analysis results with Model 2 can explain 0.58% of the variance of lifetime cannabis use in all participants of the Sister Study, achieved with p-value cutoff at 10^−5^.

### Sensitivity analysis

We conducted additional analyses to adjust for other factors that influence DNA methylation as shown in previous studies: alcohol use^28^ and BMI.^29^ We examined if the EWAS results with Model 2 differed by expanding Model 2 to also include alcohol use and body mass index (BMI) as covariates. The effect sizes of the top cannabis use–associated CpGs (*p* < 0.001) from Model 2 (Supplementary Table 1) were strongly correlated (*r* = 0.99) with the results of the sensitivity analyses (Supplementary Fig. 5).

### Follow-up of CpGs significantly associated with lifetime cannabis use

We examined the five CpGs that were epigenome-wide significantly associated with lifetime cannabis use, four of which were identified using Model 2 in all participants (adjusted for cigarette smoking) and one CpG identified in participants who never smoked cigarettes. Using the EWAS Atlas^30^ (https://ngdc.cncb.ac.cn/ewas/tools), we found that three of the five CpGs were significantly correlated with nearby gene expression in brain (Supplementary Table 4, *cor*(cg01101459, *LINC*01132) = 0.121, *cor*(cg00813162, *ACTN*1) = −0.135, *cor*(cg14237301, *APOBR*) = −0.389)).

We also looked up the correlation of DNAm for the five CpGs between whole blood and brain regions (prefrontal cortex, entorhinal cortex, superior temporal gyrus, and cerebellum) through an existing database (http://epigenetics.essex.ac.uk/bloodbrain/).^31^ Among the five CpGs, DNAm at cg22572071-*ADGRF1* and cg14237301-*APOBR* showed moderate correlations between whole blood and prefrontal cortex (*r* > 0.2). At cg22572071-*ADGRF1* and cg15280358-*ADAM12*, DNAm was moderately correlated (*r* > 0.2) between whole blood and cerebellum (Supplementary Fig. 6, Supplementary Table 5).

Enrichment analyses comparing the top cannabis use–associated CpGs from Model 2 to previously reported EWAS results in EWAS Atlas identified associations with other diseases and environment exposures (Supplementary Table 6). Among the enriched traits detected, Crohn’s disease, alcohol consumption, BMI, and multiple sclerosis were significantly overlapped with our EWAS results.

Interestingly, cannabis use–associated CpGs identified in participants who never smoked cigarettes showed significant overlap with CpGs previously associated with smoking (*p* = 5.58 × 10^−8^), smoking cessation, lung function, and lung carcinoma.

To explore whether the DNAm changes were genetic-driven, we looked up the methylation quantitative trait loci (meQTLs) for the five epigenome-wide significant CpGs in the Genetics of DNA Methylation Consortium (GoDMC) database^32^ (http://mqtldb.godmc.org.uk/about), and overlapped them with the GWAS results for lifetime cannabis use^27^ (Supplementary Table 7). None of the meQTLs were significantly associated with lifetime cannabis use (*p* = 5 × 10^−8^), indicating that our significantly associated CpGs were not directly driven by the genetic variants. The meQTLs for cg00813162-*ACTN1* and cg14237301-*APOBR* showed some degree of significance in the GWAS (*p* < 0.01, Supplementary Table 7).

### Differentially methylated regions

We used an R software tool, ipDMR,^33^ to identify DMRs based on EWAS meta-analysis results (Supplementary Table 8). Model 1 (no adjustment for cigarette smoking) yielded 514 significant DMRs (FDR < 0.05) with a minimum of two probes, while Model 2 (accounting for cigarette smoking) had 10 significant DMRs, showing that one of the four epigenome-wide significant CpGs (cg00813162-*ACTN1*) reside in a region where proximate CpGs were associated with lifetime cannabis use. Additionally, there were nine DMRs that included no single epigenome-wide significant CpG, and instead multiple correlated CpGs showed some evidence of association. The DMR analysis in participants who never smoked cigarettes identified six DMRs with a minimum of two probes, none of which included the epigenome-wide significant CpG sites. The gene set enrichment analysis of the genes that overlap with the DMRs from Model 2 identified four Gene Ontology (GO) biological processes (Supplementary Fig. 7): growth, response to metal ion, actin filament bundle organization, and cellular response to zinc ion.

## Discussion

In this study, we performed the largest EWAS meta-analyses of lifetime cannabis use to date (9,436 multi-ancestry participants) using DNAm data from peripheral blood samples. The basic model showed that DNAm changes associated with lifetime cannabis use largely overlapped with cigarette smoking– associated DNAm sites. After additionally adjusting for smoking in the EWAS model, we found four CpG sites statistically independent of cigarette smoking that were significantly associated with lifetime cannabis use. Additionally, we conducted EWAS in participants who never smoked cigarettes to further eliminate the influence of the participants’ cigarette smoking. This analysis showed high consistency with the smoking-adjusted model in all participants (Model 2) and yielded one additional CpG significantly associated with lifetime cannabis use. The genes annotated to these five cannabis use–associated CpGs are relevant to a range of health outcomes.^34-44^

The five epigenome-wide significant CpGs are annotated to the nearest genes: *LINC01132, ADGRF1, ADAM12, ACTN1*, and *APOBR*. Of these, cg01101459-*LINC01132*, cg00813162-*ACTN1*, and cg14237301-*APOBR* were inversely associated with cannabis use, meaning that individuals who had used cannabis had lower DNAm at these CpG sites compared to those who had never used cannabis. *LINC01132* is a long noncoding RNA gene that has been reported to function as an oncogene that relates to the malignant behaviors of cancer cells.^34,45,46^ Lower expression of *LINC01132* has been associated with reduced oncogenicity. *ACTN1* (Alpha-Actinin-1) encodes a non-muscle cytoskeletal protein that binds actin to the cell membrane.^35^ Genetic variants and differential expression of the *ACTN1* have been reported in various diseases, including congenital macrothrombocytopenia, Angelman syndrome, Bowen disease, postmenopausal osteoporosis, lupus erythematosus, and COVID-19.^35-38,47,48^ *APOBR* (Apolipoprotein B receptor) encodes a receptor protein that binds to dietary triglyceride-rich lipoproteins. Its genetic variants have been associated with obesity,^39,49^ bladder cancer,^42^ pneumonia,^41^ allergy,^40^ and lifetime cannabis use.^27^ The results from our analyses using EWAS Atlas showed evidence that the expression of *LINC01132* in brain is positively correlated with the DNAm level of cg01101459, while the expression of *ACTN1* and *APOBR* in the brain are negatively correlated with the DNAm levels of the corresponding CpGs. However, both DNAm and gene expression patterns are tissue-specific, and their correlations merit further investigation in different tissues. Genetic variants in *APOBR* have been linked to lifetime cannabis use in GWAS.^27^ However, we did not find overlap between the meQTLs and significant GWAS SNPs. Future EWAS integrating meQTLs is needed to confirm whether the association is genetic-driven.

On the other side, cg22572071-*ADGRF1* and cg15280358-*ADAM12* were positively associated with cannabis use. *ADGRF1* is a receptor gene that that is critical in neurodevelopment and neuroinflammation.^43,50^ The overexpression of *ADGRF1* has been reported in breast cancer.^51^ *ADAM12* encodes protein that involving in cell-cell interaction, muscle development, and neurogenesis. The expression of *ADAM12* has been reported to be upregulated in various tumor cells and is an emerging prognostic biomarker for cancer.^52-56^ Genetic variants in *ADAM12* have been associated with neurological diseases such as multiple sclerosis and Alzheimer’s disease. Further experimental studies are needed to determine the potential effects of changes in DNAm levels on the corresponding gene expressions.

The majority of significant CpGs in EWAS for cannabis use with the basic model (Model 1) overlapped with those identified in the EWAS for cigarette smoking.^26^ The number of significant CpGs decreased dramatically after adjusting for smoking status in our Model 2. These findings suggest that cigarette smoking is a strong confounder for cannabis use. However, we found that DNAm scores calculated as weighted sums of the beta values of significant CpGs from Model 1 explained a greater amount of variance in lifetime cannabis use than the DNAm scores based on Model 2 (3.79% vs. 0.58%). Even in participants who never smoked cigarettes, the DNAm scores based on Model 1 could explain 0.91% of the variance in lifetime cannabis use. Additionally, the EWAS results in participants who never smoked cigarettes showed enrichment of CpGs associated with cigarette smoking. These results suggest that cannabis use and cigarette smoking may independently influence the DNAm levels of these CpGs.

In previous studies, DNAm scores have been used to predict various outcomes with the variance explained ranging from 0.6% in low-density lipoprotein, 2.5% in educational attainment, 12.5% in alcohol use, to 60.9% in cigarette smoking.^57^ The variance in lifetime cannabis use explained by DNAm scores was (3.79%) is moderate. Future applications that integrate both polygenic risk scores and DNAm scores may improve prediction power.^57,58^ The accumulation of even larger scale GWAS and EWAS for lifetime cannabis use will be necessary to establish reliable biomarkers for clinical purposes.

As a complement to the main EWAS, the DMR analysis that combined nearby correlated CpGs identified additional regions associated with cannabis use. After adjusting for smoking, the EWAS for lifetime cannabis use identified fewer epigenome-wide significant hits, and the DMR analysis provides an improved power to detect correlated CpGs with small effects influenced by cannabis exposure. The gene set enrichment analysis revealed biological processes related to growth, response to metal ion and zinc ion, and assembly of actin filament bundle.

The large sample set included in this study provides a good representation for a wide range of populations across countries, sexes, ancestry groups, and age ranges, empowering more generalizable findings in identifying a common and robust DNAm signature for lifetime cannabis use. A recent genetic study^59^ has shown that increased sample sizes of diverse ancestries improved detection power and generated more generalizable polygenic risk scores. However, one limitation of this approach is that associations with ancestry effects may have been attenuated when all data were combined, as heterogeneity across different cohorts would have reduced power to detect such specific associations in underrepresented populations. Future studies on more data from diverse populations may reveal ancestry-, sex- and age-specific DNAm associations. There may also be relevant confounders that we have not been able to adjust for, such as exposures and experiences that lead to cannabis use.

In this study, we analyzed DNAm profiles from blood samples. While DNAm profiles differ across tissues and cell types, our results based on blood samples may not be generalizable to other tissues that may be more biologically relevant to the addiction and other behavioral effects of cannabis use, such as the brain. Using data from an online database, we found moderate positive correlations between DNAm in whole blood and prefrontal cortex or cerebellum at three of the five cannabis use–associated CpG sites. Further experimental validation is needed to connect the DNAm changes in blood and other tissues that may relate to health effects associated with cannabis use. It should also be noted that the current EWAS results may suffer from potential confounding effects caused by blood cell subtype heterogeneity that were not fully captured by reference-based deconvolution that we applied.^60,61^

The EWAS meta-analysis conducted in this study only reflects the association between lifetime cannabis use and DNAm levels, without any causal inference. More precise measurements of cannabis use regarding recency and frequency are needed to further investigate the persistent and transient effects on DNAm changes. Taken together, we propose future studies to integrate cannabis use patterns, GWAS results, DNAm data from multiple tissues, and gene expression data, to infer causal links between cannabis exposure and DNAm levels.

In conclusion, our EWAS found that a large proportion of DNAm changes that are significantly associated with cannabis use overlap with those observed for cigarette smoking. After adjusting for smoking status in the EWAS and conducting additional association testing in participants who never smoked cigarettes, we identified five cigarette smoking-independent CpGs that were significantly associated with lifetime cannabis use. Three of these CpGs showed significant correlations with the expression of nearby genes that have been linked to various health outcomes. These findings provide insights into DNAm profiles that are shared between smoking and cannabis use or specific to each substance, and suggest a substantial proportion of the variance in lifetime cannabis use are captured by DNAm. Follow-up studies are warranted to unravel the biological relevance of the differential DNAm to health outcomes.

## Methods

### Study cohorts

This study included data from seven participating cohorts: the Sister Study,^62^ Gulf Long-Term Follow-Up Study (GuLF),^63^ Netherlands Twin Register (NTR),^64^ Veteran Aging Cohort Study (VACS),^65^ Finnish Twin Cohort (FinnTwin),^66^ Avon Longitudinal Study of Parents and Children (ALSPAC),^67^ and UK Adult Twin Registry (TwinsUK).^68^ The final sample size consisted of 9,436 participants, including 4,190 individuals who reported ever using cannabis and 5,246 who reported never using cannabis (Table 1). Detailed information for each cohort can be found in the Supplementary Methods. Informed consent was obtained from each participant, and each study was approved by their Institutional Review Boards.

### Cannabis assessment

Our analyses focused on lifetime cannabis use based on self- or parent-report. Participants were classified as ever users if they reported using cannabis at least once prior to the blood sample collection used to generate DNAm data, and as never users if they reported never using cannabis prior to the blood draw. This definition of the phenotype aligns with the “Substances—Lifetime Use” variable in the PhenX Toolkit,^69^ making the results comparable and combinable.

### DNA methylation measurements

DNAm was measured in peripheral blood using either the Illumina Infinium HumanMethylation450 BeadChip (450K array, 76%) or the Illumina Infinium Methylation EPIC BeadChip (EPIC array, 24%), as shown in Table 1. DNAm levels were calculated as *β*-values, which represent the percentage of DNA that is methylated at the interrogated CpG site and ranges from 0 to 1. Quality control and normalization procedures were implemented consistently across all cohorts, with considerations specific to each cohort (detailed in the Supplementary Methods).

### EWAS for lifetime cannabis use

In each cohort, the association between DNAm levels and lifetime cannabis use was tested under a linear model or a GEE model if participants were related. We stratified the EWAS analyses by ancestry groups (EA and AA) and DNAm array types (450K and EPIC). For each CpG site, the DNAm beta value was considered as the outcome with lifetime cannabis use as the predictor of interest, and two separate models were applied. In the basic model (Model 1), we included sex (except in cohorts with only one sex), age at blood collection, blood cell type estimation, and technical covariates. In Model 2, we additionally adjusted for cigarette smoking status defined as current, former, or never.

Comparing with prior smoking EWAS,^25^ the results from Model 2 were considered as cigarette smoking– independent DNAm biomarkers for lifetime cannabis use. Additionally, we implemented EWAS within the subset of participants who never smoked cigarettes using Model 1 to minimize the possible confounding effect of cigarette smoking. More detailed information for models and covariates used in each cohort is provided in the Supplementary Methods.

### Meta-analysis

We summarized cohort- and ancestry-specific EWAS results using inverse variance fixed effects meta-analysis implemented in the METAL software,^70^ with Model 1 (all participants), Model 2 (all participants), and Model 1 in participants who never smoked cigarettes, respectively. We reported overlapping CpGs between 450K and EPIC arrays, which included 452,453 CpG probes. Epigenome-wide significance was defined as FDR less than 5%. Manhattan and QQ plots were genearted using the *CMplot* function within the R package rMVP.^71^ Heterogeneity among the studies was assessed using the Cochran’s Q-test implemented in METAL.

### Methylation scores

As the single largest cohort included in this study, the Sister Study was reserved as the testing dataset to evaluate DNAm scores. For each individual in the Sister Study, a methylation score was calculated as a weighted sum of CpGs significantly associated with lifetime cannabis use in the EWAS meta-analysis conducted without the Sister Study.^58^ At a given CpG site *i*, the methylation beta value (*meth*_*i*_) was multiplied by the effect size of the CpG in the meta-analysis (*eff*_*i*_). Then a methylation score was obtained by summing over a selected CpG set:

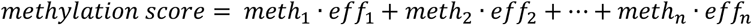

We applied different p-value thresholds to select the significant CpG sets for both Model 1 (*p* < 10^−1^, 10^−3^, 10^−5^, 10^−7^, 10^−9^, 10^−11^, 10^−13^) and model 2 (*p* < 10^−1^, 10^−3^, 10^−5^, 10^−7^). To evaluate performance of the methylation scores, we quantified the percentage of variance (*R*^2^) explained as proposed by Lee et al.^72^ for binary responses.

### Integrating EWAS results with gene expression

To investigate the potential relationship between DNAm and gene expression levels, we used the correlations between DNAm and expression data available in six tissues (brain, colon, kidney, liver, stomach, testis) in the EWAS Atlas.^30^ Specifically, we analyzed the DNAm levels of the significant CpGs to determine if they were associated with the expression levels of nearby genes.

### Correlation of DNAm between blood and brain tissues

An online database (https://epigenetics.essex.ac.uk/bloodbrain/)^31^ was used to examine the correlations of DNAm between whole blood and four brain regions (prefrontal cortex, entorhinal cortex, superior temporal gyrus, and cerebellum), respectively. For each CpG site, a boxplot was generated to display the distribution of DNAm levels across all five tissues, and the Pearson correlation was calculated between the DNAm level in whole blood and each of the four brain tissues.

### Enrichment analysis against previous EWAS results

To determine the potential overlap of our top CpGs with previously reported EWAS results, we used the EWAS Atlas toolkit^30^ to conduct enrichment analyses. To meet the minimum input requirement, we selected the top 20 CpGs from the EWAS meta-analysis using Model 2 and the top 20 CpGs in participants who never smoked cigarettes, and ran enrichment analyses on each group separately.

### Methylation quantitative trait loci (meQTL)

To explore the potential genetic basis for DNAm changes identified in our study, we looked up genetic variants that are associated with DNAm levels of CpG sites within the Genetics of DNA Methylation Consortium (GoDMC) database,^32^ which includes both local (*cis*) and distal (*trans*) meQTLs. We examined the overlap between the meQTL variants for cannabis use–associated CpG sites and the variants identified in a GWAS for lifetime cannabis use.^27^

### DMR analyses

We used an R software tool, ipDMR,^33^ to identify DMRs in which a cluster of correlated CpGs showed evidence for association with lifetime cannabis use. ipDMR calculates an overall p-value for small intervals bordered by two adjacent CpGs based on the association p-values from an EWAS analysis. It then combines all nearby significant intervals (using a seed threshold) and calculates an FDR-adjusted p-value for the combined region. We applied ipDMR to summary statistics from our EWAS meta-analysis with the following parameters: seed p-value < 0.05, maximum distance to combine adjacent intervals 1000bp and bin size 50bp. To assess the biological significance of the identified DMRs, we then conducted gene set enrichment analysis using the tool provided by Functional Mapping and Annotation of Genome-Wide Association Studies (FUMA)^73^ to test for enrichment of the genes that overlapped with the DMRs in predefined pathways.

### Sensitivity analyses

To evaluate the potential confounding effects of alcohol use and BMI on the association between DNA methylation and lifetime cannabis use, we conducted sensitivity analyses. In these analyses. We compared the effect sizes and p-values of the significant CpGs identified in the main analysis with those obtained in the sensitivity analyses to assess the robustness of our results. Detailed definitions of alcohol use in each cohort can be found in the Supplementary Methods section.

### Comparison of results across ethnic groups

Considering the differential DNAm patterns across different ethnicity groups, we conducted a separate EWAS meta-analysis in EA (N=7,795) and AA (N=1,641) groups with Model 2. We compared the effect sizes and p-values of the top significant CpGs from the main EWAS results with the results in each ancestry group.

## Data Availability

All data are existing from the seven established cohorts: (1) Avon Longitudinal Study of Parents and Children (ALSPAC), (2) Sister Study, (3) Gulf Long-Term Follow-Up (GuLF) Study, (4) Veterans Aging Cohort Study (VACS), (5) Netherlands Twin Register (NTR), (6) Finnish Twin Cohort (FinnTwin), and (7) U.K. Adult Twin Registry (TwinsUK). Data access can be requested for three of the cohorts (ALSPAC, Sister Study, and GuLF Study) by submitting an application outlining the proposed use of the data. Upon approval by the study's Executive Committee and payment of a data access charge, data will be shared with investigators. DNA methylation array and genotype array data from two of the cohorts VACS will be deposited to NCBI's Gene Expression Omnibus (GEO) and the Database of Genotypes and Phenotypes (dbGaP), respectively. The parent studies that funded the original data generation will be responsible for depositing the data and making them publicly available in compliance with the National Institutes of Health (NIH) Genomic Data Sharing policy. The remaining three cohorts (NTR, FinnTwin, and TwinsUK) are international and do not follow the same data sharing policies as the NIH-funded U.S. cohorts. However, collaborations with study investigators, as shown here, may be possible.

## Acknowledgements

This work was mainly supported by National Institute on Drug Abuse grant number R01DA048824 (PI: Fang). We also acknowledge the contributions of the staff and participants of all cohorts involved in this study, and cohort-specific funding and acknowledgement are included in Supplementary Section.

## Author contributions

All authors reviewed the results and approved the manuscript. F.F., B.Q., D.B.H., and E.O.J. conceived and designed the study. F.F., B.Q., K.G.L., J.vD., J.A.M., S.L., M.L., V.V.O., R.C., Z.X., L.Z., M.M., and Y.X. conducted cohort-specific analyses. F.F, J.A.M., and L.Z. performed meta-analysis. M.O., J.A.T., J.T.B., J.K., D.I.B., K.X., and D.P.S. oversaw the cohort research team and contributed to study design, analysis plan and interpretation of the results. J.M.V. and L.J.B. provided consulting to study design, analysis plan and interpretation of the results.

