## Supplemental information and figures for "Trans-ancestry epigenome-wide association meta-analysis of DNA methylation with lifetime cannabis use"

**Supplementary Methods**

**Description of participating cohorts**

**ALSPAC Study**: ALSPAC stands for Avon Longitudinal Study of Parents and Children, which is a UK birth cohort study to examine the health of 14,500 families in the area around Bristol.^1,2^ DNA methylation (DNAm) was examined for DNA extracted from blood of mothers during antenatal and two follow-up periods using HumanMethylation450 BeadChip. For children, DNAm was examined in cord blood and blood in follow-up periods when they were approximately 7, 15, and 17 years old. For the phenotype in our study, only child data collected in the last two follow-up periods were used. To assess lifetime cannabis use, we investigated all the questionnaires before blood collection; if the participant answered “yes” to the question “Respondent has tried cannabis” in any time before the blood collection date, we counted it as ever used. Smoking and drinking were coded as never, ever, and current using the longitudinal data before the time of DNA extraction. Please note that the study website contains details of all the data available through a fully searchable data dictionary and variable search tool and reference the following webpage: <http://www.bristol.ac.uk/alspac/researchers/our-data/>. The cell type proportions were estimated using the Houseman method^3^ implemented in in the R package minfi.^4^ Body mass index (kg/m2) was computed based on weight and height obtained at the same time point as DNA extraction. Surrogate variables obtained by applying surrogate variable analysis were included to adjust for unknown technical artefacts such as batch effects.

The epigenome-wide association study (EWAS) between DNAm and lifetime cannabis use was conducted under a linear model with DNAm $\beta$-values as outcome. Besides lifetime cannabis use, Model 1 included the following predictors: age at DNA extraction, sex, cell type proportions, and surrogate variables. Model 2 was additionally adjusted for cigarette smoking.

Ethical approval for the study was obtained from the ALSPAC Ethics and Law Committee and the Local Research Ethics Committees. Informed consent for the use of data collected via questionnaires and clinics was obtained from participants following the recommendations of the ALSPAC Ethics and Law Committee at the time. Consent for biological samples was collected in accordance with the Human Tissue Act (2004).

**FinnTwin Study**: The Finnish Twin Cohort Study include three longitudinal cohorts,^5^ the Older Twin cohort,^6^ FinnTwin16 (FT16),^7^ and FinnTwin12 (FT12).^8^ Here, we studied twins belonging to the FT16 and FT12, which are longitudinal studies of five consecutive birth cohorts (born in 1975–1979, n = 2,800 pairs, and 1983–1987, n = 2,700, respectively) of Finnish monozygotic and dizygotic twins who have completed surveys and interviews beginning in adolescence and continuing into adulthood (FT16 at age 16, 17, 18, 24, 34; FT12 at age 11, 14, 17.5, 22). Lifetime cannabis use was assessed in the intensive samples of FT12 (mean age 22) and FT16 (mean age 26) through Semi-Structured Assessment for Genetics of Alcohol (SSAGA).^9^ Twins answering “yes” to the question “Have you ever used cannabis?” were coded as ever users. Tobacco smoking and alcohol use were both coded as current, former, and never. BMI was based on measured height and weight. All the phenotypic variables were collected at the same research visit with the blood draw. Altogether 1,429 twins had cannabis use and whole blood DNA methylation data available and were included in the current study.

DNA was extracted from whole blood with standard procedures, and bisulfite conversion was performed using the EZ-96 DNA MethylationGold Kit (Zymo Research Corp, Irvine, CA, USA) following the manufacturer’s instructions. DNAm was quantified using the Infinium HumanMethylation450 or EPIC BeadChip Kit (Illumina, San Diego, CA, USA) according to the manufacturer’s protocol. Methylation data for 450k and EPIC data was preprocessed using R package meffil.^10^ First, beta values were background corrected and then functional normalization was performed, using 13 PCs for the 450k dataset and 17 PCs for the EPIC dataset. Probes with a detection p-value > 0.05 or a beadcount < 3 in greater than 10% of samples were removed. We additionally removed probes on sex chromosomes and those known to be cross-reactive. We estimated cell types using the FlowSorted.Blood.EPIC reference set. Beta-mixture quantile normalization was used to adjust beta values for differences because of probe type.^11^ After QC, 1,368 samples with good quality 450K or EPIC data remained for analysis.

The association between cannabis use and DNA methylation was tested under a generalized estimation equation (GEE) model with DNA methylation β-value as outcome. All models included the following predictors: cannabis use, sex, age at blood sampling, percentages of CD8T, CD4T, natural killer cells, and neutrophils, and bisulfite conversion sample plate. EWAS analyses were performed with the R package “gee” with zygosity and family to account for relatedness of the samples.

Informed consent was obtained from all participants. All study procedures were approved by the ethics committees of Helsinki University Central Hospital (113/E3/2001, 249/E5/2001, 346/E0/05, and 270/13/03/01/2008).

**GuLF Study**: The GuLF Study is a prospective U.S. cohort designed to examine health effects of the Deepwater Horizon oil spill disaster.^12^ The analytic samples included in this study were men, aged 21-65 years, who participated in oil spill cleanup effort. The sample selection prioritized those with more biological samples (e.g., urine or serum) and more follow-up data. In total, DNAm of 1,584 participants were measured with Illumina Infinium MethylationEPIC Beadchip using whole blood samples. A total of 1,555 samples for 834183 CpGs passed QC: detection P value <1x10-6, number of beads ≥3, outlier samples or CpGs based on the distribution of total intensity plots, and Bisulfite conversion efficiency > 4000, CpG coverage per sample or per CpG> 0.95%. We estimated cell type proportions with the Houseman method^3^ and surrogate variables to adjust for unknown technical artefacts implemented in the R package Enmix.^13^ The following preprocessing steps were performed: (1) Background correction using ENmix method, (2) dye bias correction using RELIC method, (3) inter-array quantile normalization, and (4) probe type bias adjustment using Regression on Correlated Probes (RCP) method. For each CpG the outlier data points (beta value outside of 3IRQ range from mean) were excluded; missing values were imputed using KNN method.

Lifetime cannabis use was assessed during the clinical exam by the question “Ever used marijuana.” Smoking and alcohol use were both coded as never, former, and current. Body mass index (kg/m2) was computed based on weight and height obtained at the same time point as blood drawn.

EWAS was run with a linear model using DNAm $\beta$-values as outcome. Besides lifetime cannabis use, Model 1 included following predictors: age at DNA drawn, Sample plate, cell type proportions, and surrogate variables. Model 2 additionally adjusted for cigarette smoking.

Written informed consent was obtained from all participants at a home visit. The study protocol was reviewed by the Institute of Medicine in September 2010 and was approved by the Institutional Review Board of the NIEHS. The study is overseen by a Scientific Advisory Board and a Community Advisory Board.

**NTR**: The Netherlands Twin Register is a population-based cohort of over 200,000 people from across the Netherlands. It consists of twin-families (i.e., twins, their parents, spouses, and siblings aged between 0 and 99 years at recruitment) and started around 1987 with newborn twins and adolescent and adult twins. Full details have been reported previously.^14^ DNA was collected from buccal cells and whole blood as part of multiple projects. For the current paper, we analyzed DNA methylation measured in whole blood collected in the NTR-Biobank study.^15-17^ Good quality whole blood DNA methylation data were available for 3,087 samples from 3,055 individuals, including monozygotic and dizygotic twins and parents, siblings, and spouses of twins. The current EWAS included individuals with information on cannabis use and covariates (Model 1, N = 2,142 samples; Model 2, N = 2,109 samples; participants who never smoked cigarettes, N = 1,253).

Information on cannabis use was obtained in longitudinal NTR surveys. For the current project, the following questions on cannabis initiation from NTR surveys 8 and 10 were used: From survey 8, the question “Did you ever experiment with hash marijuana?” If participants answered this question with yes, they were asked to fill out age at first use. From survey 10, the questions “Have you ever used cannabis? We are referring to hash, weed, marijuana, a joint, or space cake.” and “How old were you when you first used cannabis?” Participants who reported that they had initiated cannabis use before blood draw were classified as ever users, and participants who had not initiated cannabis use before blood draw were classified as never users. Information on current and past smoking behavior was also collected as part of the NTR biobank project at the moment of blood draw. Smoking was coded according to the analysis plan as: 0 = never smoker, 1 = former smoker, 2 = current light smoker (1-10 cigarettes per day), 3 = current moderate smoker (11-19 cigarettes per day), 4 = current heavy smoker (>= 20 cigarettes per day).

DNA methylation was assessed with the Infinium HumanMethylation450 BeadChip Kit (Illumina, San Diego, CA, USA) by the Human Genotyping facility (HugeF) of ErasmusMC, the Netherlands (<http://www.glimdna.org/>) as part of the Biobank-based Integrative Omics Study (BIOS) consortium.^18^ DNA methylation measurements have been described previously.^17,18^ Genomic DNA (500ng) from whole blood was bisulfite treated using the Zymo EZ DNA Methylation kit (Zymo Research Corp, Irvine, CA, USA), and 4 µl of bisulfite-converted DNA was measured on the Illumina 450k array following the manufacturer’s protocol. A number of sample- and probe-level quality checks and sample identity checks were performed, as described in detail previously.^17^ In short, sample-level QC was performed using MethylAid.^19^ Probes were set to missing in a sample if they had an intensity value of exactly zero, or a detection p > .01, or a bead count of < 3. After these steps, probes that failed based on the above criteria in > 5% of the samples were excluded from all samples (only probes with a success rate ≥ 0.95 were retained). The following probes were also removed: sex chromosomes, probes with a single nucleotide polymorphism within the CpG site (at the C or G position) irrespective of minor allele frequency in the Genome of the Netherlands population, irrespective of minor allele frequency,^20^ and ambiguous mapping probes reported by Chen et al. with an overlap of at least 47 bases per probe.^21^ The methylation data were normalized with functional normalization.^22^

The association between cannabis use and DNA methylation was tested under a GEE model with DNA methylation β-value as outcome. All models included the following predictors: cannabis use, sex, age at blood sampling, percentages of monocytes, eosinophils, and neutrophils, HM450k array row, and 96-wells bisulfite sample plate (dummy coding). EWAS analyses were performed with the R package “gee.” The following settings were used: Gaussian link function (for continuous data), 100 iterations, and the “exchangeable” option to account for the correlation structure within families and within persons.

Informed consent was obtained from all participants. The study was approved by the Central Ethics Committee on Research Involving Human Subjects of the VU University Medical Centre, Amsterdam, an Institutional Review Board certified by the U.S. Office of Human Research Protections (IRB number IRB00002991 under Federal-wide Assurance- FWA00017598; IRB/institute codes, NTR 03-180).

**Sister Study**: Sister Study is a U.S. nationwide prospective cohort of women with a sister who has breast cancer.^23^ The participants were women aged 35-75 years with no personal history of breast cancer at baseline. DNAm was examined from whole blood samples collected along with questionnaires for a random sample set of 1,336 women and 1,542 additional women who later developed in situ or invasive breast cancer during follow-up. All women included in this study were clinically breast cancer free at the time of the blood draw used for DNAm data generation with HumanMethylation450 BeadChip.

In total, DNAm of 1,584 participants were measured with Illumina Infinium MethylationEPIC Beadchip using whole blood samples. A total of 1,555 samples for 834183 CpGs passed QC: detection P value <1x10-6, number of beads ≥3, outlier samples or CpGs based on the distribution of total intensity plots, and Bisulfite conversion efficiency > 4000, CpG coverage per sample or per CpG > 0.95%. We estimated cell type proportions with the Houseman method^3^ and surrogate variables to adjust for unknown technical artefacts implemented in the R package Enmix.^13^ The following preprocessing steps were performed: (1) Background correction using ENmix method, (2) dye bias correction using RELIC method, (3) inter-array quantile normalization, and (4) probe type bias adjustment using RCP method. For each CpG the outlier data points (beta value outside of 3IRQ range from mean) were excluded, missing values were imputed using KNN method.

The lifetime cannabis use was assessed by response to the question: “Have you ever smoked marijuana?” Smoking was coded as never, ever, and current. Alcohol use was coded as current and non-current. Body mass index (kg/m2) was computed based on weight and height obtained at the same time point as blood draw.

EWAS was run with a linear model using DNAm $\beta$-values as outcome. Besides lifetime cannabis use, Model 1 included following predictors: age at DNA drawn, incident breast cancer status (binary), sample plate, cell type proportions, and surrogate variables. Model 2 additionally adjusted for cigarette smoking.

Written informed consent were obtained from all participants during blood sample collection at a home visit. The study was approved by the Institutional Review Boards of the National Institute of Environmental Health Sciences, National Institutes of Health, and the Copernicus Group.

**TwinsUK**: The TwinsUK registry includes over 14,000 research volunteer twin participants from the United Kingdom recruited since 1992.^24^ Volunteers are either monozygotic or dizygotic same-sex twins, predominantly female (82%), middle-aged (mean age 59) and over 18 years old. Most volunteers are healthy individuals and of European descent. Data on volunteers are collected through longitudinal questionnaires and clinical visits. In 2009 TwinsUK participants were asked whether they tried cannabis at least once in their lifetime (ever users). Ever users with DNA methylation data after/on year of questionnaire were set as cases, and never users with DNA methylation data before the questionnaire were set as controls. There were altogether 30 female ever users and 80 female never users with whole blood DNA methylation data in the TwinsUK cohort (n = 110 females). The whole blood DNA methylation data used in this study were profiled with the Illumina EPIC platform. Habitual cigarette smoking data were available for all participants of this study and habitual alcohol intake data were available for 80% of participants. Participants were characterized as never-smokers (“0”), ex-smokers (“1”) or current smokers (“2”) according to their smoking habits, and never/former drinkers (0 g/day, “0”), light drinkers (< 14 g/day, “1”), moderate drinkers (14–28 g/day, “2”) or heavy drinkers (> 28 g/day, “3”) according to their weekly consumption of alcoholic beverages. Age at time of phlebotomy and BMI were available for all participants included in the analysis.

Ethical approval was granted by the National Research Ethics Service London-Westminster, the St Thomas’ Hospital Research Ethics Committee (EC04/015 and 07/H0802/84). All twins provided written informed consent prior to taking part in research activities.

**VACS**: The Veterans Aging Cohort Study (VACS) is a prospective cohort study of veterans focusing on the clinical outcomes of HIV infection.^25^ Demographic and clinic data were obtained from the patients after they provided written consent; data were collected via telephone interviews, focus groups, and full access to the national Veterans Affairs electronic medical record system. All subjects in this subset of the VACS cohort were African American men. DNA samples were extracted from whole blood. DNA methylation was profiled by using Infinium Human Methylation 450K BeadChip (HM450K) or the Infinium Human Methylation EPIC BeadChip.^26^

The study was approved by the committee of the Human Research Subject Protection at Yale University and the Institutional Research Board Committee of the Connecticut Veteran Healthcare System. All subjects provided written consents.

**Funding and Acknowledgements**

**ALSPAC Study**: The UK Medical Research Council and Wellcome (Grant ref: 217065/Z/19/Z) and the University of Bristol provide core support for ALSPAC. This publication is the work of the authors and Fang Fang will serve as guarantors for the contents of this paper. A comprehensive list of grants funding is available on the ALSPAC website (<http://www.bristol.ac.uk/alspac/external/documents/grant-acknowledgements.pdf>). The Accessible Resource for Integrated Epigenomics Studies, which generated DNA methylation profiles was funded by the UK Biotechnology and Biological Sciences Research Council (BB/I025751/1 and BB/I025263/1). Additional epigenetic profiling on the ALSPAC cohort was supported by the UK Medical Research Council Integrative Epidemiology Unit and the University of Bristol (MC_UU_12013_1, MC_UU_12013_2, MC_UU_12013_5, and MC_UU_12013_8), the Wellcome Trust (WT088806) and the United States National Institute of Diabetes and Digestive and Kidney Diseases (R01 DK10324). We are extremely grateful to all the families who took part in this study, the midwives for their help in recruiting them, and the whole ALSPAC team, which includes interviewers, computer and laboratory technicians, clerical workers, research scientists, volunteers, managers, receptionists, and nurses.

**FinnTwin Study**: Data collection of the Finnish twin cohort samples has been supported by the Academy of Finland Center of Excellence in Complex Disease Genetics (Grants 213506 and 129680 and 336823), the Academy of Finland (Grants 265240 and 263278 to JK, and 297908 to MO), NIH Grant DA12854 (to PAFM), and by the Sigrid Juselius Foundation (to JK and MO).

**GuLF Study**: The GuLF Long-Term Follow-up Study was supported by the National Institutes of Health Common Fund and the Intramural Research Program of the National Institutes of Health, National Institute of Environmental Health Sciences (ZO1 ES 102945).

**NTR**: We acknowledge funding from the National Institute on Drug Abuse grant number R01DA048824 (PI: Fang), the Netherlands Organization for Scientific Research (NWO): Biobanking and Biomolecular Research Infrastructure (BBMRI–NL, NWO 184.033.111) and the BBRMI-NL-financed BIOS Consortium (NWO 184.021.007), NWO Large Scale infrastructures X-Omics (184.034.019), Genotype/phenotype database for behavior genetic and genetic epidemiological studies (ZonMw Middelgroot 911-09-032); Netherlands Twin Registry Repository: researching the interplay between genome and environment (NWO-Groot 480-15-001/674); the Avera Institute, Sioux Falls (USA) and the National Institutes of Health (NIH R01 HD042157-01A1, MH081802, Grand Opportunity grants 1RC2 MH089951 and 1RC2 MH089995); epigenetic data were generated at the Human Genomics Facility (HuGe-F) at ErasmusMC Rotterdam. DIB acknowledges the Royal Netherlands Academy of Science Professor Award (PAH/6635).

**Sister Study**: Sister Study was supported by the Intramural Research Program of the National Institutes of Health, National Institute of Environmental Health Sciences (Z01 ES044005, Z01 ES049033).

**VACS**: This work was supported by National Institute on Drug Abuse grant number R01DA048824 (PI: Fang); R01 DA038632 (Xu); R01 DA047063 (MPI: Xu and Aouizerat); R01 DA047820 (MPI:Xu and Aouizerat). COMpAAAS/Veterans Aging Cohort Study, a CHAART Cooperative Agreement, is supported by the National Institutes of Health: National Institute on Alcohol Abuse and Alcoholism (U24-AA020794, U01-AA020790, U01-AA020795, U01-AA020799; U10-AA013566-completed) and in kind by the U.S. Department of Veterans Affairs. Additional grant support from the National Institute on Drug Abuse R01-DA035616 is also acknowledged. The views and opinions expressed in this manuscript are those of the authors and do not necessarily represent those of the Department of Veterans Affairs or the U.S. government. This work uses data provided by patients and collected by VA as part of its care and support.

**TwinsUK**: This work was supported by the UK Economic and Social Research Council and Biotechnology and Biological Sciences Research Council (BBSRC) (ES/N000404/1 to JTB) and JPI ERA HDHL DIMENSION (BBSRC BB/S020845/1 to JTB). The TwinsUK study is funded by the Wellcome Trust, Medical Research Council, Versus Arthritis, European Union Horizon 2020, Chronic Disease Research Foundation, Zoe Global Ltd and the National Institute for Health Research Clinical Research Network and Biomedical Research Centre based at Guy’s and St Thomas’ NHS Foundation Trust in partnership with King’s College London.

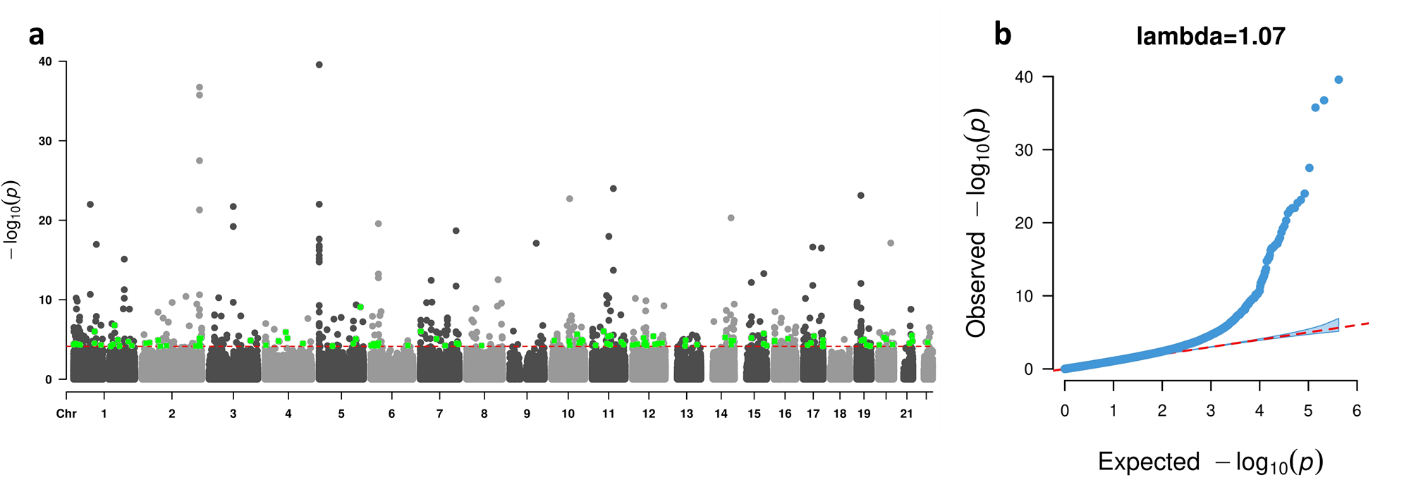

**Supplementary Figure 1:** Manhattan (**a**) and quantile-quantile (QQ) plots (**b**) from the EWAS-meta analysis for lifetime cannabis use with model 1. The dotted red line in (a) indicates the epigenome-wide significance cutoff at FDR<0.05 ($P<7.26\times{10}^{-5}$). The green dots in (a) are CpG probes that were significantly associated with cannabis use in our study and not previously reported to be significantly associated with cigarette smoking.

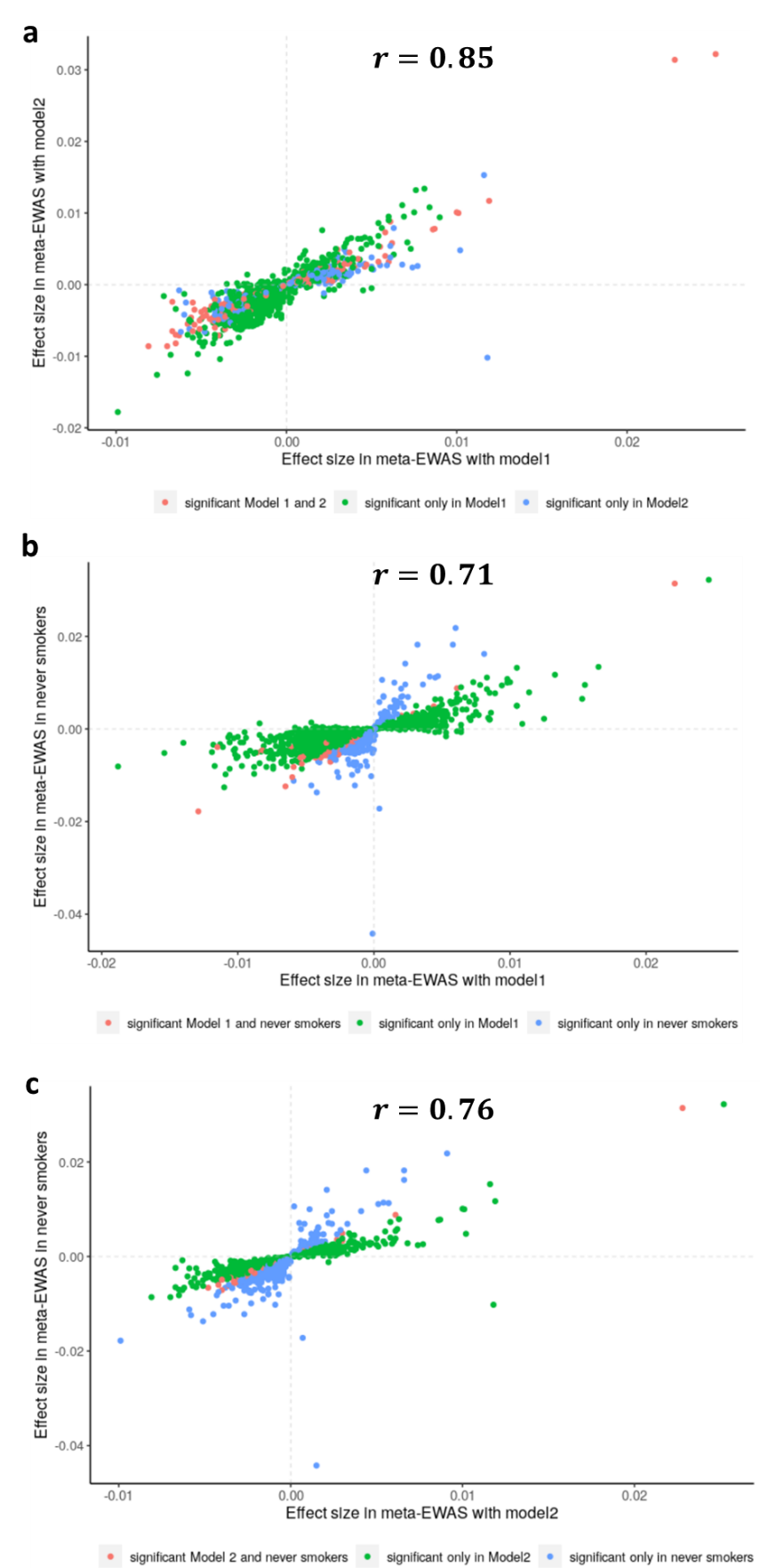

**Supplementary Figure 2:** Comparing effect sizes of top CpGs (P < 0.001) from the EWAS meta-analysis for lifetime cannabis (**a**) between Model 1 (x-axis) and Model 2 (y-axis) in all participants; (**b**) between Model 1 in all participants (x-axis) and Model 1 in participants who never smoked cigarettes (y-axis); (**c**) between Model 2 in all participants (x-axis) and Model 1 in participants who never smoked cigarettes (y-axis).

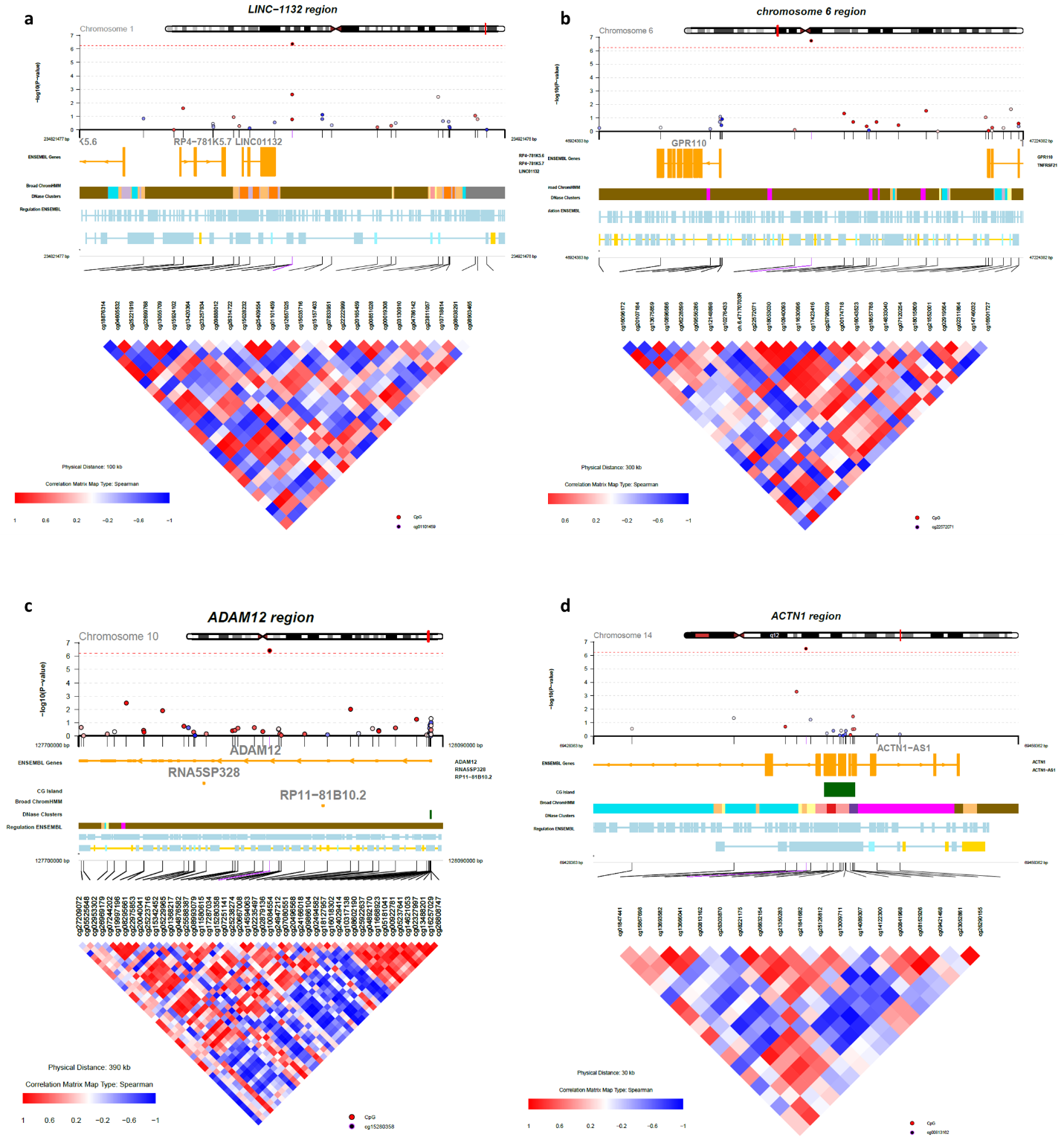

**Supplementary Figure 3:** The CoMET^27^ plots around the four CpGs significantly associated with lifetime cannabis use, independent of cigarette smoking.

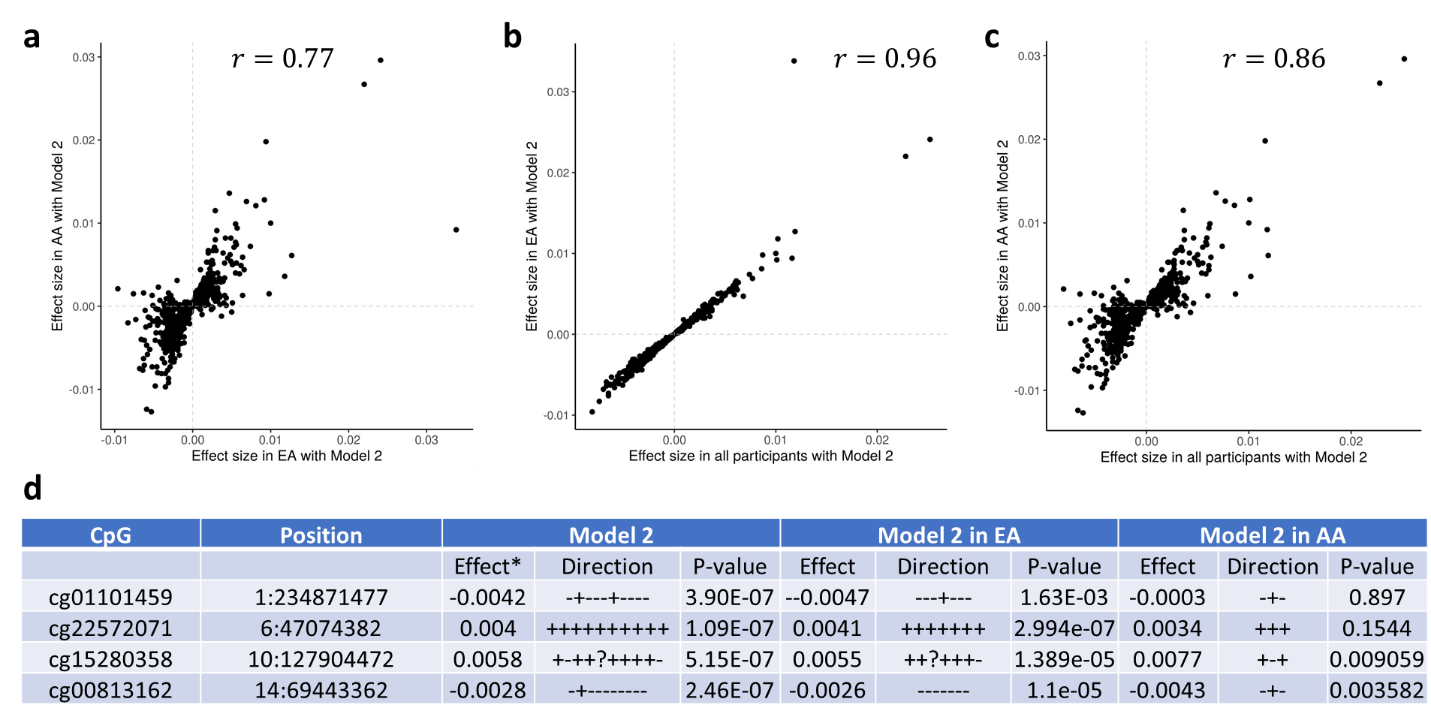

**Supplementary Figure 4:** Comparing effect sizes of top CpGs (P<0.001) in EWAS meta-analysis for lifetime cannabis use with Model 2 (x-axis) (**a**) between European ancestry (EA) and African ancestry (AA) participants; (**b**) between all and EA participants; (**c**) between all and AA participants. (**d**) All four CpGs showing epigenome-wide significance in all participants were also significant in the EA group (P < 0.05/4), and two of them were significant in the AA group.

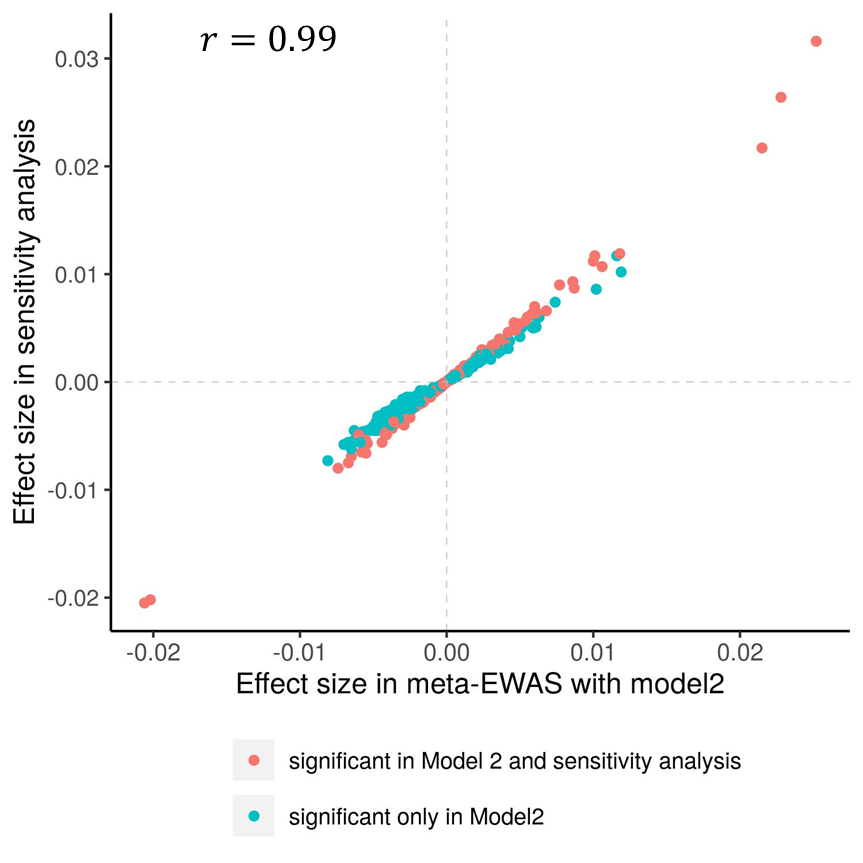

**Supplementary Figure 5.** Comparing effect sizes of top CpGs (P<0.001) in EWAS meta-analysis for lifetime cannabis use with Model 2 (x-axis) with those obtained in sensitivity analysis with additional adjustment of alcohol use and BMI (y-axis).

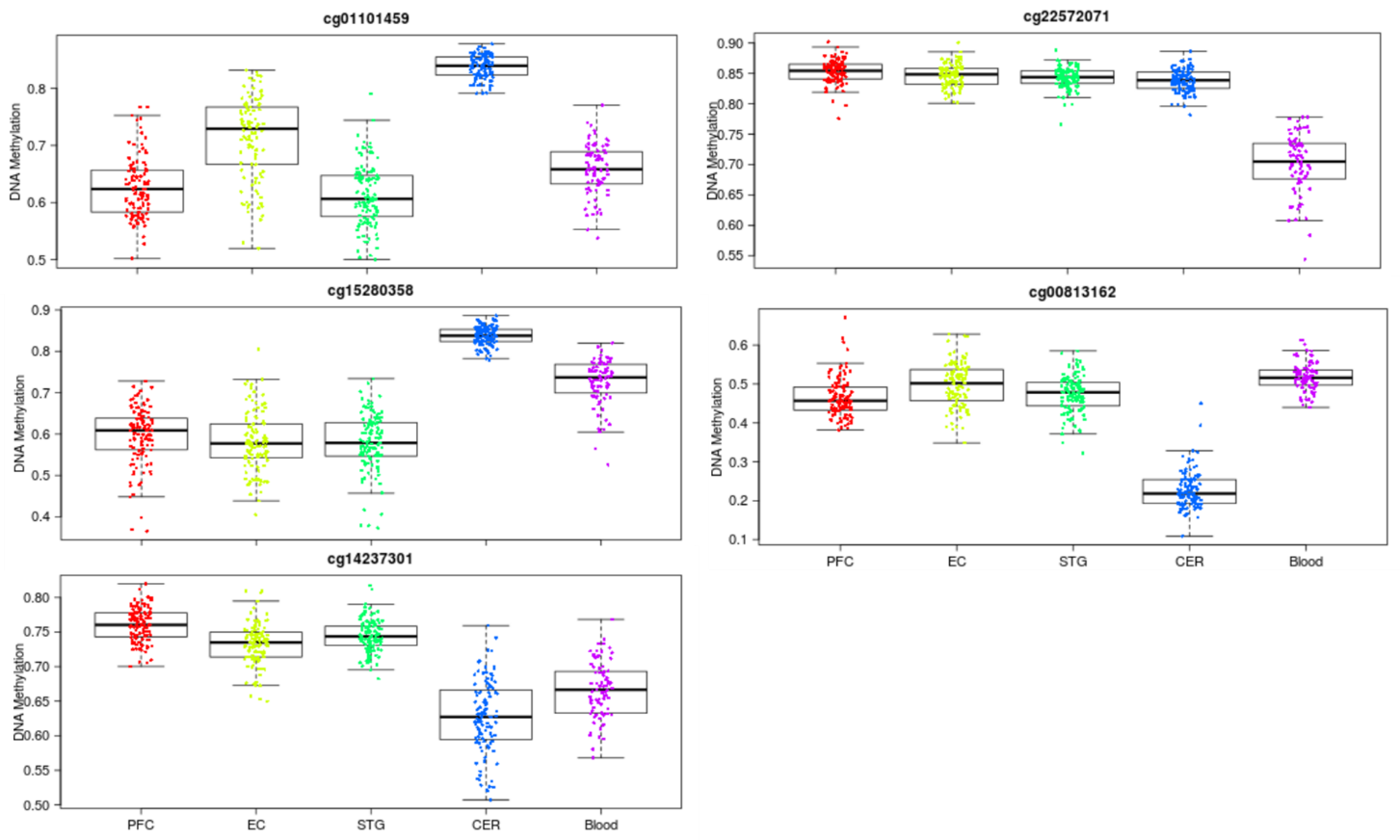

**Supplementary Figure 6:** Boxplots of the distribution of DNAm at the five CpG sites epigenome-wide significantly associated with lifetime cannabis use across tissues (blood, prefrontal cortex-PFC, entorhinal cortex-EC, superior temporal gyrus-STG and cerebellum-CER) using data from an online database (<https://epigenetics.essex.ac.uk/bloodbrain/>).

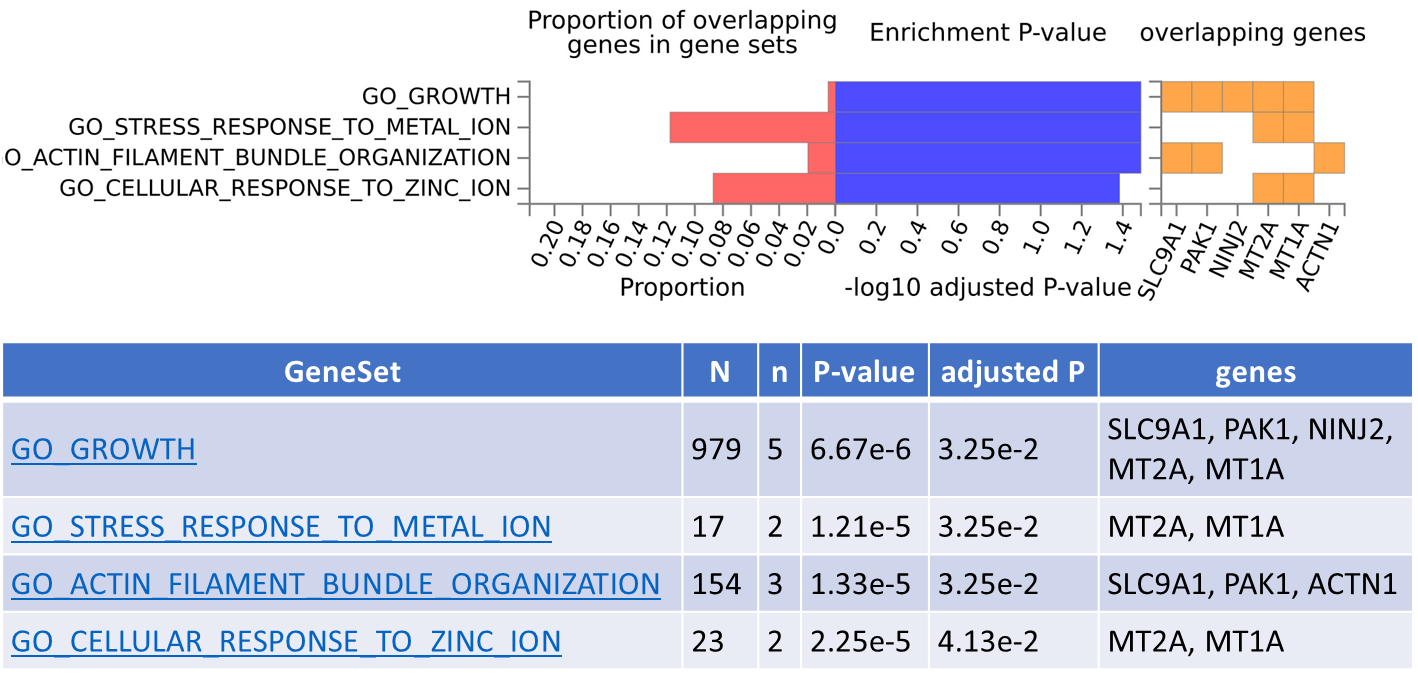
**Supplementary Figure 7:** The gene set enrichment analysis using FUMA^28^ showed four biological processes [Gene Ontology (GO)] were enriched with genes that overlapped with the DMRs detected in EWAS Model 2.
